# Organism spectrum and no-growth fraction of deep specimens in code-defined orthopedic infection: a reproducible, cross-sectional MIMIC-IV benchmark

**DOI:** 10.64898/2026.07.09.26357616

**Authors:** Yosef Adiniaev, Alon Gorenshtein, Tohar M Timor, Eyal Klang, Alexander Geftler

## Abstract

**Introduction:** Culture data guide orthopedic-infection management, yet the organism spectrum, resistance, and no-growth fraction are reported inconsistently and mostly within proprietary registries. We characterized these in a public, reproducible dataset.

**Methods:** Retrospective cross-sectional study using MIMIC-IV version 3.1, a de-identified single-center US database. Episodes with an International Classification of Diseases diagnosis of prosthetic joint infection (PJI) or native osteomyelitis were identified; organism-spectrum and no-growth analyses were restricted to the 46% with at least one deep musculoskeletal culture (tissue or bone, synovial or joint fluid, implant sonication), so the benchmark describes culture-sampled, not all, coded episodes. Proportions carry exact 95% CIs; variation was tested by logistic regression with Benjamini-Hochberg control, and an out-of-fold logistic model quantified how well no-growth was anticipated by structured data.

**Results:** Of 7697 episodes (median age, 60 years; 35.5% female), 1089 were PJI, 5715 native osteomyelitis, and 893 other device infection. Among 7700 deep specimens (3560 episodes; 2603 patients), 35.7% showed no growth (patient-clustered 95% CI, 34.0%-37.3%). The fraction was higher in PJI than osteomyelitis (48.6% vs 26.6%) but rose with sampling intensity (24.5% to 50.7%), indicating differential ascertainment. *S. aureus* led (32.5%; 43.3% methicillin-resistant), and PJI was less often polymicrobial than osteomyelitis (adjusted OR, 0.44). No-growth was weakly anticipated by structured data (out-of-fold AUROC, 0.63).

**Conclusions:** About one-third of deep specimens from code-defined orthopedic infection showed no growth. This specimen-level fraction differs from a criterion-confirmed culture-negative-infection rate and depends on sampling intensity; it is released as a re-runnable benchmark on identical open data, not a transferable rate.

## 1 Introduction

Prosthetic joint infection and native osteomyelitis are among the most consequential complications in orthopedic practice, and their management depends almost entirely on microbiological data. The choice and duration of antimicrobial therapy, the decision to retain or remove hardware, and the counseling of patients all rest on which organism is recovered, whether the infection is polymicrobial, and how resistant the isolate is (Lew and Waldvogel, 2004; Tande and Patel, 2014). A recurring clinical problem is the specimen that yields no organism at all. Culture-negative infection, reported to affect 5% to 42% of cases, forces empiric therapy and complicates source control, yet its frequency is difficult to compare across series because specimen sourcing, culture technique, and case definitions differ between institutions (Tande and Patel, 2014; Berbari et al., 2007).

Much of what is known about the microbiology of orthopedic infection comes from single-institution surgical series or from proprietary registries whose underlying records cannot be re-examined, which limits reproducibility. The no-growth fraction and organism spectrum are reported as point estimates without a shared substrate on which alternative definitions can be tested, and newer diagnostic strategies, from implant sonication to molecular assays, are evaluated against institution-specific baselines rather than an open reference. Publicly available, de-identified electronic health record data could provide such a reference but have rarely been applied to musculoskeletal infection, where the relevant specimens (bone, synovial fluid, sonicate) are a small fraction of all cultures.

We used MIMIC-IV, a large de-identified single-center database, to build an open and fully re-runnable benchmark for the microbiology of code-defined orthopedic infection. We characterized the organism spectrum, the polymicrobial fraction, and the antimicrobial-resistance profile, and quantified the no-growth fraction of deep musculoskeletal specimens with exact confidence intervals, overall and by infection type. Because a no-growth result is clinically actionable only if it cannot be anticipated in advance, we also asked how well no-growth was predicted by routinely captured structured data. The aim is descriptive and methodological: to provide a transparent reference that others can re-run on identical data, not to estimate a causal effect or a national rate.

## 2 Methods

### 2.1 Data source and reporting

We analyzed MIMIC-IV version 3.1, a de-identified database of patients admitted to a single US academic medical center, comprising hospital-wide and intensive care data and available to credentialed users under a data use agreement (Johnson et al., 2023). Reporting followed the Strengthening the Reporting of Observational Studies in Epidemiology (STROBE) and the REporting of studies Conducted using Observational Routinely-collected Data (RECORD) guidelines (**Table S 1**). Reporting of the anticipatability probe follows the Transparent Reporting of a multivariable prediction model for Individual Prognosis Or Diagnosis (TRIPOD) statement, with model details in the model card (**eMethods 6**).

### 2.2 Cohort

We identified hospital episodes (unique admissions) carrying an ICD, Ninth or Tenth Revision (ICD-9/10), diagnosis of prosthetic joint infection (infection of an internal joint prosthesis: ICD-9 996.66; ICD-10 T84.5x) or native osteomyelitis (ICD-9 730.x; ICD-10 M86.x) in any diagnosis position. Episodes coded for infection of other internal orthopedic devices (ICD-9 996.67; ICD-10 T84.6x-T84.7x), which are not joint prostheses, formed a separate device-infection group. When both prosthetic joint infection and osteomyelitis codes were present, the episode was assigned to prosthetic joint infection, with a co-occurrence flag retained for sensitivity analysis. Analyses of the organism spectrum and the no-growth fraction were restricted to episodes with at least one deep musculoskeletal culture specimen.

### 2.3 Specimen taxonomy and organism classification

Microbiology specimens were mapped from their source description to categories: deep tissue or bone, synovial or joint fluid, implant sonication or foreign body, abscess or deep fluid, superficial swab, blood, and other. The three deep musculoskeletal categories constituted the primary analytic sources; a sensitivity definition also included abscess or deep fluid. Foot cultures were classified as superficial rather than deep, because in this database they frequently represent superficial diabetic-foot specimens rather than bone or deep tissue; they contributed 66 specimens, and their reclassification did not change the benchmark. Organism names were normalized to species and higher-level groups (gram-positive, gram-negative, anaerobe, fungal, mycobacterial) with a version-controlled dictionary; low-resolution morphotype results (for example, “mixed bacterial flora,” “gram-negative rods”) were counted as growth but not as speciated isolates. The specimen and organism dictionaries are provided as **Table Ss 2** and **3**.

### 2.4 Measures

A specimen was culture-positive if it grew at least one organism and no-growth if a culture test yielded no organism. We use “no-growth” for this specimen-level measure throughout, reserving “culture-negative infection” for the distinct, criterion-confirmed clinical entity. Non-culture tests (serology, antigen, toxin, viral, and smear-only assays) were excluded. A specimen or episode was polymicrobial if at least two distinct organism species were recovered. Methicillin-resistant *S. aureus* (MRSA) was defined by an oxacillin-resistant isolate among *S. aureus* with an interpretable oxacillin result.

Resistance was reported separately for an *S. aureus* panel and a gram-negative panel rather than pooled across organisms, and, for consistency with the organism-spectrum analysis, was restricted to deep musculoskeletal isolates; agent-level resistance was the proportion of isolates reported resistant among those with an interpretable result. Diagnostic intensity was the number of distinct culture specimens and source categories per episode. In-hospital mortality, length of stay, and discharge disposition were tabulated as descriptors.

### 2.5 Statistical analysis

All proportions carry exact (Clopper-Pearson) 95% CIs. Categorical group differences were tested with the χ² test and summarized with the Cramér V effect size; count distributions were compared with the Kruskal-Wallis test and epsilon-squared. Variation in no-growth and in polymicrobial infection by age band, sex, race group, insurance category, and infection type was estimated with multivariable logistic regression, using standard errors clustered by patient and reported as adjusted odds ratios (ORs) with 95% CIs. Because MIMIC-IV lacks measured disease severity and pre-admission outpatient antibiotic exposure, these models describe variation, not causal effects. The Benjamini-Hochberg procedure controlled the false discovery rate within each variation model (2-sided *P* < .05).

To quantify how well no-growth was anticipated by routine data, we fit a logistic model (standardized features; median imputation with informative-missingness indicators) using 5-fold stratified, patient-grouped cross-validation and evaluated out-of-fold predictions by the area under the receiver operating characteristic curve (AUROC), with a 2000-sample bootstrap 95% CI and the Brier score. A clinical-context model (infection type, specimen-source mix, demographics) was compared with the same model augmented by inflammatory and nutritional markers (white blood cell count, C-reactive protein, erythrocyte sedimentation rate, albumin, neutrophil percentage) by a paired bootstrap of out-of-fold predictions. Specimen count was excluded from the feature set because it is definitionally linked to the episode-level outcome. This component is a measurement probe, not a deployable clinical tool, and was not externally validated. Analyses used Python 3.14 (pandas 3.0, scikit-learn 1.9, statsmodels 0.14, scipy 1.17) with fixed random seeds, and the analysis code is released in full.

## 3 Results

### 3.1 Cohort

Of 7697 orthopedic-infection episodes, 5715 (74.3%) were native osteomyelitis, 1089 (14.1%) prosthetic joint infection, and 893 (11.6%) other device infection (**Table 1**). The median age was 60 years, 35.5% were female, and in-hospital mortality was 2.7%. At least one deep musculoskeletal culture specimen was obtained in 3560 episodes (46.3%), more often in prosthetic joint infection (68.6%) than osteomyelitis (41.1%); these 3560 episodes (2603 patients), with 7700 deep specimens, formed the analytic cohort.

**Table 1.** Characteristics of orthopedic-infection episodes, overall and by infection.

| Characteristic | Overall | Prosthetic joint<br>infection | Native<br>osteomyelitis | Other device<br>infection |
| --- | --- | --- | --- | --- |
| No. of episodes | 7697 | 1089 | 5715 | 893 |
| Age, median (IQR), y | 60.0 (50.0-70.0) | 64.0 (55.0-73.0) | 59.0 (50.0-69.0) | 56.0 (45.0-67.0) |
| Female, % | 35.5 | 46.4 | 32.4 | 41.5 |
| White race, % | 72.5 | 76.3 | 70.6 | 80.3 |
| Black race, % | 14.7 | 10.9 | 16.6 | 6.9 |
| Medicare, % | 54.2 | 64.6 | 53.7 | 45.4 |
| Medicaid, % | 19.5 | 16.3 | 19.9 | 20.6 |
| Private insurance, % | 23.2 | 17.4 | 23.3 | 29.2 |
| ICU stay, % | 15.8 | 17.4 | 16.1 | 12.0 |
| Length of stay,<br>median (IQR), d | 7.0 (4.3-11.7) | 7.0 (4.7-11.0) | 7.0 (4.1-11.8) | 6.9 (4.3-11.7) |
| In-hospital mortality,<br>% | 2.7 | 2.9 | 2.9 | 1.7 |
| Deep MSK culture<br>obtained, % | 46.3 | 68.6 | 41.1 | 52.1 |

Abbreviations: ICU, intensive care unit; IQR, interquartile range; MSK, musculoskeletal.

### 3.2 Organism spectrum and polymicrobial infection

Among 7090 speciated isolates from deep musculoskeletal specimens, *S. aureus* was most frequent (32.5%) and coagulase-negative staphylococci second (14.3%); the full distribution is shown in Table 2 and Figure 1. The omnibus association between the organism distribution and infection type was small (Cramér V = 0.07; *P* < .001), but the difference mattered for individual pathogens: staphylococci and *Cutibacterium* were more common in prosthetic joint infection, whereas anaerobes and *Pseudomonas* were more common in osteomyelitis, a pattern relevant to empiric coverage. Among 2797 culture-positive episodes, 42.7% (95% CI, 40.9%-44.6%) were polymicrobial. Polymicrobial infection was less common in prosthetic joint infection than osteomyelitis (27.4% vs 47.7%; Cramér V = 0.18; *P* < .001), a difference that persisted after adjustment (adjusted OR, 0.44; 95% CI, 0.35-0.55).

**Table 2.** Organism distribution among deep musculoskeletal isolates, overall and by infection type.

| Organism group | Overall, No. (%) | PJI, % | Osteomyelitis, % |
| --- | --- | --- | --- |
| Staphylococcus aureus | 2305 (32.5) | 36.4 | 30.2 |
| Coagulase-negative staphylococci | 1011 (14.3) | 17.3 | 12.4 |
| Other gram-negative | 801 (11.3) | 11.4 | 10.9 |
| Streptococcus | 685 (9.7) | 10.3 | 10.6 |
| Enterococcus | 563 (7.9) | 7.6 | 8.5 |
| Other gram-positive | 528 (7.4) | 4.5 | 8.4 |
| Pseudomonas aeruginosa | 357 (5.0) | 2.6 | 6.2 |
| Anaerobe | 286 (4.0) | 0.7 | 5.6 |
| Escherichia coli | 228 (3.2) | 3.8 | 3.0 |
| Fungi/yeast | 219 (3.1) | 3.0 | 3.3 |
| Cutibacterium (Propionibacterium) | 81 (1.1) | 2.0 | 0.6 |
| Mycobacteria | 25 (0.4) | 0.2 | 0.4 |
| Other or unclassified | 1 (0.0) | 0.0 | 0.0 |

Percentages are of speciated isolates (n = 7090 overall). Low-resolution morphotype results are excluded from speciated counts (**Table S 4**). The Anaerobe row excludes Cutibacterium (Propionibacterium), which is reported as a separate row. PJI, prosthetic joint infection.

**Figure 1.**
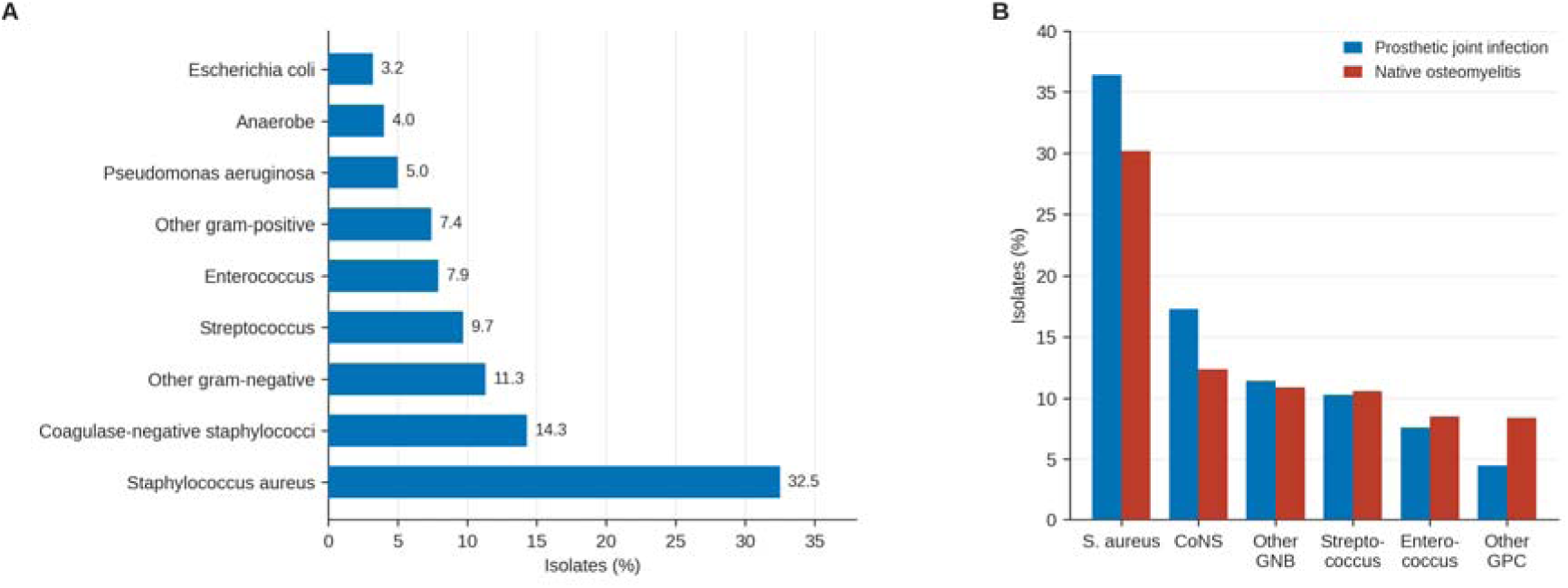
Organism spectrum of deep musculoskeletal isolates. (A) Distribution of the leading organism groups among 7090 speciated isolates. (B) The six most frequent groups compared between prosthetic joint infection and native osteomyelitis. Bars show the percentage of isolates. CoNS, coagulase-negative staphylococci; GNB, gram-negative bacilli; GPC, gram-positive cocci.

### 3.3 No-growth benchmark

Among the 7700 deep musculoskeletal specimens, 35.7% showed no growth (naive 95% CI, 34.6%-36.7%; patient-clustered bootstrap 95% CI, 34.0%-37.3%). Because specimens cluster within episodes and patients, the patient-clustered interval is the appropriate one, and specimen-level subgroup intervals are naive and understate uncertainty. At the episode level, 21.4% (95% CI, 20.1%-22.8%) of episodes with a deep specimen had no organism recovered from any deep specimen. No-growth was higher in prosthetic joint infection than osteomyelitis (48.6% vs 26.6%; Cramér V = 0.23) and varied by specimen source, being highest for synovial or joint-fluid specimens and lowest for implant sonication (**Figure 2A-2B**). It rose monotonically with the number of specimens obtained per episode, from 24.5% with a single deep specimen to 50.7% with seven or more (**Figure 2C; Table S 5**). This gradient is consistent with more intensive sampling raising the measured no-growth fraction, although the direction of a cross-sectional association is not identified, because early negative cultures may themselves prompt further sampling. Restricting to primary-position diagnoses, which selects the most clinically definite and typically more heavily sampled cases, raised the fraction to 44.6% (**Table S 6**); excluding the 97 dual-coded episodes rather than assigning them to prosthetic joint infection left the fraction (35.1%) and the infection-type contrast (48.4% vs 26.6%) essentially unchanged.

**Figure 2.**
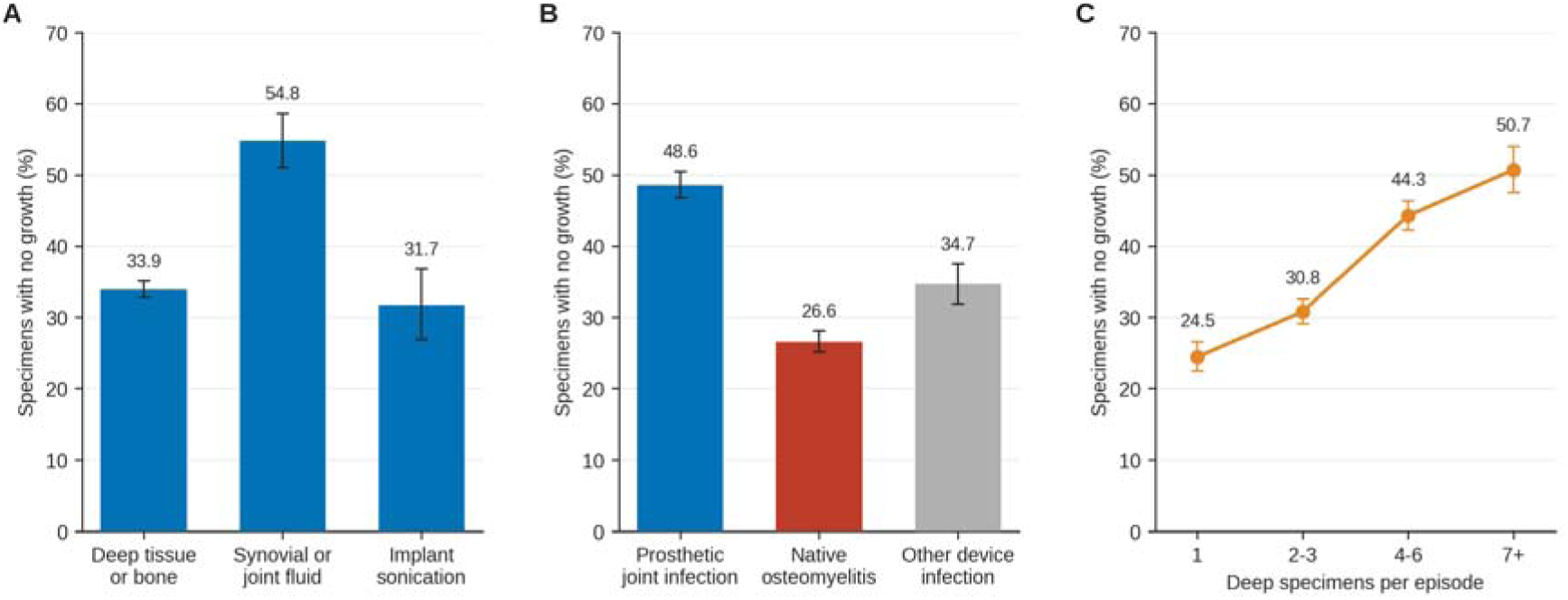
No-growth fraction of deep musculoskeletal culture specimens. No-growth fraction (A) by specimen source, (B) by infection type, and (C) by the number of deep specimens obtained per episode. Error bars are exact 95% CIs. The rise across panel C shows that more intensive sampling raises the measured no-growth fraction, which accounts for part of the prosthetic joint infection versus osteomyelitis difference in panel B.

### 3.4 Antimicrobial resistance

Resistance is reported for an *S. aureus* panel and a pooled gram-negative panel, restricted to deep musculoskeletal isolates (**Table 3**; **Figure 3**). Methicillin resistance was present in 43.3% (95% CI, 40.4%-46.2%) of deep-specimen *S. aureus* with an oxacillin result (n = 1143), similar in prosthetic joint infection (43.4%) and osteomyelitis (43.7%); no isolate was vancomycin-resistant (0 of 509). In the gram-negative panel, resistance was highest to ciprofloxacin and lowest to carbapenems, which does not capture intrinsic resistance in non-fermenters such as *Stenotrophomonas*. Susceptibility was tested only against the agents the laboratory selected, so the denominator is given per agent (**Table S 7**).

**Table 3.** Antimicrobial resistance among deep musculoskeletal isolates, by organism. Resistance is the percentage of deep-specimen isolates reported resistant among those with an interpretable susceptibility result, reported separately for each organism group so that intrinsic resistance is not conflated across organisms.

| Antimicrobial | Isolates tested | Resistant, % (95% CI) |
| --- | --- | --- |
| Oxacillin | 1143 | 43.3 (40.4-46.2) |
| Erythromycin | 1141 | 61.6 (58.7-64.4) |
| Levofloxacin | 1134 | 39.4 (36.6-42.3) |
| Clindamycin | 1081 | 38.9 (36.0-41.9) |
| Tetracycline | 1022 | 8.8 (7.1-10.7) |
| Trimethoprim-sulfamethoxazole | 1143 | 3.5 (2.5-4.7) |
| Rifampin | 570 | 2.8 (1.6-4.5) |
| Gentamicin | 1143 | 2.4 (1.6-3.4) |
| Vancomycin | 509 | 0.0 (0.0-0.7) |

*Gram-negative bacilli*
| Antimicrobial | Isolates tested | Resistant, % (95% CI) |
| --- | --- | --- |
| Ciprofloxacin | 876 | 24.9 (22.1-27.9) |
| Trimethoprim-sulfamethoxazole | 677 | 23.0 (19.9-26.4) |
| Ceftriaxone | 615 | 19.5 (16.5-22.9) |
| Cefepime | 870 | 11.1 (9.1-13.4) |
| Gentamicin | 875 | 9.7 (7.8-11.9) |
| Piperacillin-tazobactam | 819 | 7.8 (6.1-9.9) |
| Tobramycin | 870 | 7.1 (5.5-9.0) |
| Meropenem | 863 | 4.6 (3.3-6.3) |

**Figure 3.**
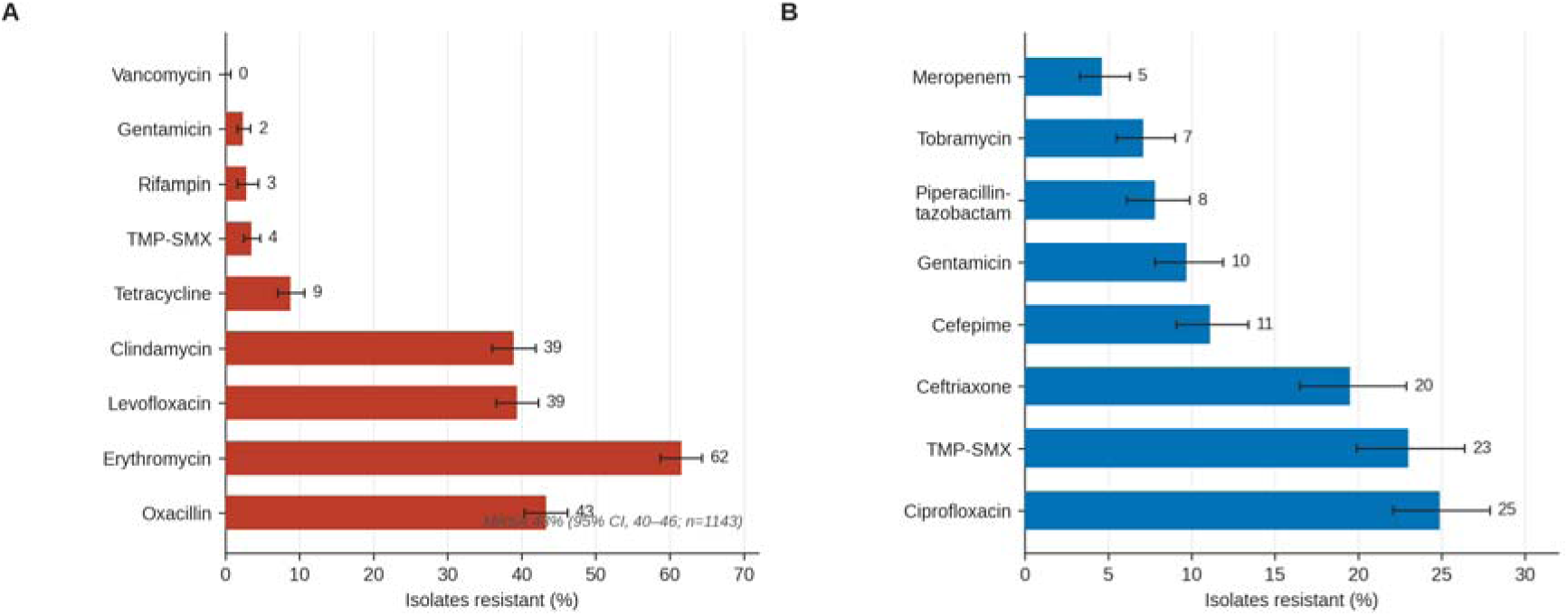
Antimicrobial resistance among deep musculoskeletal isolates, by organism. Percentage of isolates reported resistant for each agent among those with a susceptibility result, shown separately for (A) *Staphylococcus aureus* and (B) gram-negative bacilli. Error bars are exact 95% CIs; the methicillin-resistant fraction of *S. aureus* is annotated in panel A. TMP-SMX, trimethoprim-sulfamethoxazole.

### 3.5 Diagnostic intensity and yield

Episodes underwent a median of 4 (IQR, 2-7) culture specimens across a median of 2 (IQR, 1-3) source categories, more in prosthetic joint infection (median, 6) than osteomyelitis (median, 3) (epsilon-squared = 0.03; *P* < .001). Among sterile-site specimens, deep tissue or bone and implant sonication recovered organisms most often, whereas blood cultures rarely did (**Figure 4A**). Superficial swabs, excluded from the deep-specimen analyses, were positive more often, but swab positivity reflects colonization and contamination, not diagnostic yield.

**Figure 4.**
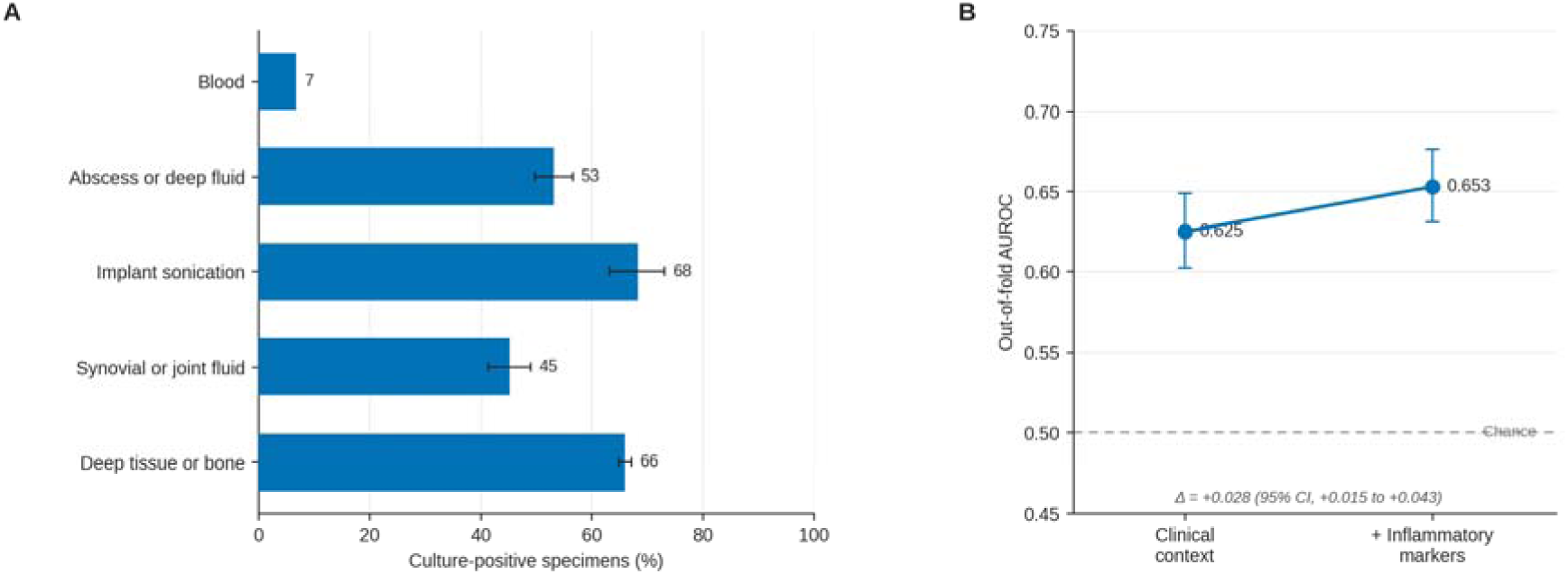
Diagnostic yield and anticipatability of no-growth. (A) Culture-positive fraction by deep and systemic specimen source, with 95% CIs; superficial swabs are excluded because their positivity reflects colonization and contamination rather than diagnostic yield. (B) Out-of-fold AUROC for anticipating no-growth from clinical context alone and with inflammatory and nutritional markers added; the dashed line marks chance discrimination. AUROC, area under the receiver operating characteristic curve.

### 3.6 Variation and anticipatability

In logistic models with patient-clustered standard errors, no-growth was associated with prosthetic joint infection (OR, 1.84; 95% CI, 1.52-2.22) and female sex (OR, 1.43; 95% CI, 1.21-1.70), both retained after false-discovery-rate control, but not with age band, race group, or insurance category (all adjusted *P* > .05); polymicrobial infection showed no sociodemographic association. These models omit specimen count by design, so the prosthetic-joint-infection odds ratio reflects the same sampling-confounded contrast rather than an independent biological effect. No-growth was only weakly anticipated by routine data: a clinical-context model achieved an out-of-fold AUROC of 0.625 (95% CI, 0.602-0.649), and adding inflammatory and nutritional markers changed discrimination minimally (0.653; paired Δ = 0.028) (**Figure 4B**). The result held under alternative handling of the missing markers, and in the small complete-case subsample (n = 323) discrimination was no better than chance (AUROC, 0.48). Weak discrimination is consistent with no-growth being driven by factors absent from the record, such as pre-admission antibiotics, but does not establish this, because a simple model may be underspecified.

## 4 Discussion

In open, reproducible data, the organism spectrum, resistance profile, and specimen-level no-growth fraction of code-defined orthopedic infection could be characterized transparently and re-run by others. About one-third of deep specimens (35.7%) grew no organism, a fraction that tracked sampling intensity more than infection type and that routine structured data, including inflammatory markers, did not anticipate.

These results extend prior work, with one caveat about what is being measured. Published no-growth fractions are typically computed among cases meeting a formal infection definition, such as the 2018 International Consensus Meeting criteria (Parvizi et al., 2018) or the European Bone and Joint Infection Society criteria (McNally et al., 2021). The present quantity is instead the fraction of deep specimens with no growth among code-defined episodes; these related but distinct estimands make the reported 5% to 42% range a context, not a direct comparator. Prior antimicrobial exposure and fastidious or low-burden organisms are recognized contributors (Berbari et al., 2007; Osmon et al., 2013). The higher no-growth fraction of synovial or joint-fluid specimens is consistent with the lower sensitivity of fluid aspirates relative to tissue and sonicate sampling, and possibly with aspirates being obtained after antibiotics have started; both underlie guidance to obtain multiple periprosthetic tissue specimens and, where available, implant sonication (Osmon et al., 2013; Trampuz et al., 2007). The predominance of staphylococci and the greater polymicrobial burden of osteomyelitis are concordant with surgical series (Lew and Waldvogel, 2004; Tande and Patel, 2014).

The higher no-growth fraction in prosthetic joint infection than osteomyelitis should not be read as biological. Deep cultures were obtained in a far larger share of prosthetic-joint-infection episodes, because revision arthroplasty routinely triggers multiple protocolized intraoperative specimens, including in patients with low pretest probability, whereas osteomyelitis is often cultured only once infection is clinically evident. Because no-growth rose steeply with sampling intensity, the more heavily sampled prosthetic-joint-infection episodes show a higher fraction partly for that reason rather than through biology. Conditioning the cohort on a deep culture having been obtained, which itself depends on infection type and clinical suspicion, is a collider-type selection that can distort the internal contrast, not only limit generalizability. We therefore present the contrast descriptively; separating the mechanisms would require linkage to procedure context or explicit selection modeling. The association of female sex with no-growth (OR, 1.43), adjusted for infection type but not for sampling intensity, remains unexplained and should be read as hypothesis-generating rather than as a biological difference.

Two implications follow. First, because the measured no-growth fraction depends on how intensively episodes are sampled, institutional culture-negative or no-growth rates should not be compared across centers without matching sampling intensity; such a rate is interpretable only alongside the specimen-per-episode distribution that produced it. Second, because no-growth was weakly anticipated even with inflammatory markers, a fraction that could instead be predicted from documentation would more likely signal a correctable workflow artifact than true sterility, and should not be treated as noise to be adjusted away.

### 4.1 Limitations

The main strength of this study is transparency: by reproducing established microbiological patterns in open data and releasing the analysis, it provides a shared substrate on which case definitions, specimen taxonomies, and newer diagnostics can be compared without a proprietary registry. Several limitations bound interpretation. The cohort was defined by administrative codes, which are imperfect markers of true infection; we therefore framed it as code-defined and counted organisms at the isolate level without an organism-significance rule, so low-virulence single isolates count as pathogens. MIMIC-IV is a single academic center enriched for intensive care, so the organism and resistance profiles, including the high methicillin-resistance fraction, may not generalize; the contribution is a reproducible reference, not a national estimate.

Agent-level resistance is measured only among the isolates the laboratory chose to test, which may overestimate resistance where testing was prompted by treatment failure or organism complexity. Only in-hospital records are captured, excluding outpatient antibiotic administration, a recognized driver of no-growth. The analytic cohort was restricted to the 46% of episodes with a deep culture, a selection that can bias both generalizability and the internal infection-type contrast (**Table S 8**). Finally, the reported quantity is a specimen-level no-growth fraction, related to but not identical with the criterion-confirmed culture-negative-infection rate.

## 5 Conclusions

Approximately one-third of deep orthopedic-infection specimens showed no growth in open, reproducible data, with a no-growth fraction and organism profile that differed between prosthetic joint infection and native osteomyelitis and were not readily explained by routine structured variables. Because the fraction depends on how intensively episodes are sampled, its value is a transparent, re-runnable reference on identical open data and a method others can apply to their own records, rather than a target rate for direct cross-institution comparison, which would require matched sampling intensity.

## Supporting information

STROBE-RECORD reporting checklist

## Data Availability

The data underlying this study are the MIMIC-IV v3.1 database, available to credentialed users from PhysioNet (https://physionet.org/content/mimiciv/3.1/) under the PhysioNet Credentialed Health Data Use Agreement. The authors had no special access privileges. All analysis code required to reproduce the results is publicly available at https://github.com/yosefad1305-arch/culture-negative-ortho-mimic-iv and archived at Zenodo (https://doi.org/10.5281/zenodo.21268251).

## Code availability

All analysis code required to reproduce the results is publicly available at https://github.com/yosefad1305-arch/culture-negative-ortho-mimic-iv and permanently archived at Zenodo (Adiniaev et al., 2026; https://doi.org/10.5281/zenodo.21268251). The code runs against MIMIC-IV version 3.1, obtained independently by each user under the PhysioNet Credentialed Health Data Use Agreement; no patient-level data are distributed with the code.

## Data availability

The data are the Medical Information Mart for Intensive Care IV (MIMIC-IV) version 3.1, a credentialed, de-identified database available to approved users from PhysioNet (https://physionet.org/content/mimiciv/3.1/) under the PhysioNet Credentialed Health Data Use Agreement, after completion of the required human-subjects research training. No patient-level data are included in this article or in the accompanying code.

## Author contribution

Y.A. contributed to conceptualization, methodology, software, formal analysis, data curation, visualization, and writing of the original draft. A.Gor. contributed to methodology, validation, and review and editing of the manuscript. T.M.T. contributed to investigation, data curation, and review and editing of the manuscript. E.K. contributed to conceptualization, supervision, and review and editing of the manuscript. A.Gef. contributed to validation, resources, supervision, and review and editing of the manuscript. All authors reviewed and approved the final manuscript and agree to be accountable for all aspects of the work.

## Ethical statement

This study was a secondary analysis of MIMIC-IV version 3.1, a de-identified database of patients admitted to the Beth Israel Deaconess Medical Center, accessed under the PhysioNet Credentialed Health Data Use Agreement after completion of the required human-subjects research training. The collection of the data and creation of the resource were reviewed by the Institutional Review Board of the Beth Israel Deaconess Medical Center (Boston, Massachusetts, USA), which granted a waiver of informed consent and approved the data-sharing initiative; the Institutional Review Board of the Beth Israel Deaconess Medical Center waived ethical approval for this secondary analysis. Because all data are de-identified and were analyzed anonymously, no additional institutional review board approval or patient consent was required, consistent with the Declaration of Helsinki. No identifiable patient information was accessed, and no patient-level data are reported.

## Acknowledgements

During the preparation of this work, the authors used a large language model (Claude, Anthropic) to assist with manuscript editing, code review, and formatting of tables and figures. All study design, analyses, interpretations, and conclusions are the authors’ own. The authors reviewed and verified all content and take full responsibility for the integrity of the work.

