## Supplementary material for "Organism spectrum and no-growth fraction of deep specimens in code-defined orthopedic infection: a reproducible, cross-sectional MIMIC-IV benchmark"

### Supplementary Information

This Supplement contains the eMethods, the specimen and organism dictionaries, the full organism and susceptibility tables, sensitivity analyses, and the model card for the anticipatability probe.

#### eMethods

##### 1. Data source

MIMIC-IV version 3.1 is a de-identified relational database derived from the electronic health record of a single US academic medical center, covering admissions from 2008 to 2022, released by the Laboratory for Computational Physiology and distributed through PhysioNet under a credentialed data use agreement. Dates are independently shifted per patient into the future to preserve de-identification, so intervals within a patient are preserved but calendar time and time of day across patients are not interpretable. This analysis used the hosp module (admissions, patients, diagnoses_icd, procedures_icd, microbiologyevents, labevents) and the icu module (icustays).

##### 2. Cohort construction

Episodes were hospital admissions (hadm_id) carrying at least one qualifying ICD-9 or ICD-10

diagnosis in any position. Diagnostic code prefixes were: prosthetic joint infection (internal

joint prosthesis), ICD-9 996.66, ICD-10 T84.5; other internal orthopedic device infection, ICD-9 996.67, ICD-10 T84.6 and T84.7; native osteomyelitis, ICD-9 730, ICD-10 M86. Episodes with both prosthetic joint infection and osteomyelitis codes (n = 97) were assigned to prosthetic joint infection for the primary contrast and analyzed separately in sensitivity testing. The analytic cohort for the organism-spectrum and no-growth aims was restricted to episodes with at least one deep musculoskeletal culture specimen (n = 3560; the culture-sampled subpopulation, 46% of coded episodes).

##### 3. Specimen and organism classification

Specimens were classified from spec_type_desc into source categories (Table S 2). The three deep

musculoskeletal categories (deep tissue or bone; synovial or joint fluid; implant sonication or foreign body) formed the primary analytic set. A culture test was identified from test_name (any bacterial or fungal culture; serology, antigen, toxin, viral, and smear-only assays were excluded). Organism names (org_name) were normalized with a version-controlled dictionary (Table S 3) to species and to broad groups (gram-positive, gram-negative, anaerobe, fungal, mycobacterial). Administrative and viral-antigen strings were treated as non-organisms. Low-resolution morphotype results (for example, "mixed bacterial flora," "gram-negative rods") were counted as growth for the culture-positive determination but were not counted as speciated isolates and did not contribute to the polymicrobial species count.

##### 4. Measure definitions

Measure definitions (culture-positive, no-growth, polymicrobial, methicillin-resistant *S. aureus*, and diagnostic intensity) are as in the main-text Methods. Agent-level resistance was computed among isolates with an interpretable susceptibility result (susceptible, intermediate, or resistant), with only "resistant" counted in the numerator.

##### 5. Statistical detail

Proportions carry exact Clopper-Pearson 95% CIs (statsmodels proportion_confint, method="beta").

Categorical differences used the χ² test with the Cramér V effect size; count distributions used the

Kruskal-Wallis test with epsilon-squared = (H − k + 1)/(n − k). Multivariable logistic regression

(statsmodels) estimated adjusted odds ratios with 95% CIs for no-growth and polymicrobial

infection, with covariates age band, sex, race group, insurance category, and infection type. The

Benjamini-Hochberg procedure controlled the false discovery rate within each variation model's

covariate family (the no-growth model and the polymicrobial model, corrected separately); logistic models used cluster-robust (sandwich) standard errors on subject_id. Omnibus chi-square and Kruskal-Wallis tests are reported descriptively.

##### 6. Model card (anticipatability probe)

| Field | Value |
| --- | --- |
| Purpose | Measurement probe: how well is deep-specimen no-growth anticipated from routine structured data (not a deployable tool) |
| Outcome | Episode-level deep-specimen no-growth (all deep specimens no growth) |
| Cohort | Episodes with >=1 deep MSK culture (n = 3560; 21.4% episode-level no-growth) |
| Model M1 | Logistic regression; features: infection type, deep source-category flags, age, sex, race group, insurance |
| Model M2 | M1 + white blood cell count, C-reactive protein, erythrocyte sedimentation rate, albumin, neutrophil percentage (log-transformed where skewed; informative-missingness indicators) |
| Preprocessing | Median imputation, standardization (StandardScaler) |
| Validation | 5-fold stratified, patient-grouped cross-validation on subject_id (no patient split across folds); CV seed 42, bootstrap seed 20260702 |
| Metrics | AUROC with 2000-sample bootstrap 95% CI; Brier score; paired bootstrap for the M2 − M1 difference |
| Result | M1 AUROC 0.625 (0.602-0.649); M2 AUROC 0.653 (0.631-0.676); Δ 0.028 (0.015-0.043) |
| Sensitivity (M2b) | M1 + WBC/CRP/neutrophil only (dropping ESR and albumin): AUROC 0.643 (0.619-0.665) |
| Complete-case (all markers measured, n = 323) | M1 AUROC 0.48; M2 AUROC 0.56 |
| Excluded feature | Specimen count (definitionally tied to the all-negative outcome) |
| Marker missingness | WBC 0.7%, C-reactive protein 38%, ESR 76%, albumin 57%, neutrophil % 38% |
| Software | Python 3.14; scikit-learn 1.9; statsmodels 0.14; scipy 1.17 |
| External validation | None |

#### Table S 1. STROBE/RECORD checklist

A completed STROBE checklist for cross-sectional studies, extended with the RECORD items for routinely collected health data, is provided as a separate file with the submission package (STROBE-RECORD_checklist.docx), with each item cross-referenced to the relevant section of the manuscript and this Supplement.

Routinely collected health data, is provided with the submission package. Key RECORD items: the code lists used to define the population (eMethods 2), the validation of those codes (not performed; cohort framed as code-defined), the data-cleaning and linkage steps (eMethods 2-3), and the database governance and access (eMethods 1).

#### Table S 2. Specimen source taxonomy

| Source category | spec_type_desc members (case-insensitive) | Deep MSK |
| --- | --- | --- |
| Deep tissue or bone | TISSUE, BONE, BIOPSY (BONE MARROW excluded to "other") | Yes |
| Synovial or joint fluid | JOINT FLUID, PROSTHETIC JOINT FLUID, SYNOVIAL | Yes |
| Implant sonication or foreign body | FOREIGN BODY, Sonication culture (prosthetic joint), Foreign Body - Sonication Culture | Yes |
| Abscess or deep fluid | ABSCESS, FLUID (other), Fluid received in blood-culture bottles, FLUID CULTURE | Sensitivity only |
| Superficial swab | SWAB, Staph aureus swab, FOOT CULTURE | No |
| Blood | BLOOD CULTURE (all bottle types) | No |
| Other | URINE, SPUTUM, STOOL, RESPIRATORY, screens, serology | No |

#### Table S 3. Organism dictionary (summary)

The complete rule set is version-controlled in code/org_map.py. Broad groups: gram-positive (staphylococci [*S. aureus*; coagulase-negative staphylococci], streptococci, enterococci, corynebacteria and other gram-positive rods), gram-negative (Enterobacterales and non-fermenters including *Pseudomonas*, *Acinetobacter*, *Stenotrophomonas*), anaerobes (Bacteroides, Prevotella, Clostridium, anaerobic cocci, *Cutibacterium/Propionibacterium*, which is reported as its own group given its role in culture-negative prosthetic joint infection while remaining within the anaerobe supergroup), fungi and yeast, and mycobacteria. Administrative strings

(For example, CANCELLED, POSITIVE, NEGATIVE) and viral-antigen results were classified as non-organisms.

#### Table S 4. Full organism counts

| Organism (species-level category) | Isolates, n | % of speciated isolates |
| --- | --- | --- |
| Staphylococcus aureus | 2,305 | 32.5 |
| Coagulase-negative staphylococci | 1,011 | 14.3 |
| Enterococcus spp. | 563 | 7.9 |
| Corynebacterium spp. | 467 | 6.6 |
| Pseudomonas aeruginosa | 357 | 5.0 |
| Streptococcus, group B | 354 | 5.0 |
| Escherichia coli | 228 | 3.2 |
| Enterobacter spp. | 215 | 3.0 |
| Streptococcus spp. (other) | 207 | 2.9 |
| Bacteroides spp. | 194 | 2.7 |
| Proteus spp. | 180 | 2.5 |
| Candida spp. | 152 | 2.1 |
| Klebsiella spp. | 102 | 1.4 |
| Serratia spp. | 87 | 1.2 |
| Streptococcus anginosus group | 83 | 1.2 |
| Cutibacterium acnes | 72 | 1.0 |
| Prevotella spp. | 59 | 0.8 |
| Citrobacter spp. | 51 | 0.7 |
| Other gram-positive rod/coccus (skin flora) | 45 | 0.6 |
| Acinetobacter spp. | 40 | 0.6 |
| Yeast | 39 | 0.6 |
| Streptococcus, group A | 39 | 0.6 |
| Other gram-negative bacillus | 38 | 0.5 |
| Morganella morganii | 36 | 0.5 |
| Stenotrophomonas maltophilia | 32 | 0.5 |
| Mycobacterium spp. | 25 | 0.4 |
| Other fungus | 23 | 0.3 |
| Bacillus spp. (not anthracis) | 16 | 0.2 |
| Other anaerobe | 14 | 0.2 |
| Clostridium spp. | 11 | 0.2 |
| Providencia spp. | 10 | 0.1 |
| Pseudomonas spp. (other) | 10 | 0.1 |
| Cutibacterium/Propionibacterium spp. | 9 | 0.1 |
| Anaerobic gram-positive cocci | 8 | 0.1 |
| Aspergillus spp. | 5 | 0.1 |
| Streptococcus pneumoniae | 2 | 0.0 |
| Other/unclassified organism | 1 | 0.0 |

*Speciated isolate counts among deep musculoskeletal culture specimens across all 37 species-level categories. Percentages are of all speciated isolates; low-resolution morphotype results (for example, "mixed bacterial flora") counted toward culture-positivity but are not speciated and are not listed here. The leading groups are summarized in Table 2 of the main text.*

#### Table S 5. No-growth fraction by sampling intensity

| Deep specimens per episode | No-growth, n/total | % (95% CI) |
| --- | --- | --- |
| 1 | 446/1820 | 24.5 (22.5-26.6) |
| 2-3 | 837/2716 | 30.8 (29.1-32.6) |
| 4-6 | 984/2220 | 44.3 (42.2-46.4) |
| 7 or more | 479/944 | 50.7 (47.5-54.0) |

The no-growth fraction rose monotonically with the number of deep specimens obtained. This gradient is consistent with more intensive sampling raising the measured no-growth fraction, though the direction is not identified in cross-section, because early negative cultures may themselves prompt further sampling. It contributes to the higher fraction seen in the more heavily sampled prosthetic-joint-infection episodes.

#### Table S 6. Sensitivity analyses for the no-growth fraction

| Analysis | No-growth, n/total | % (95% CI) |
| --- | --- | --- |
| Primary (deep MSK specimens; foot cultures classified as superficial) | 2746/7700 | 35.7 (34.6-36.7) |
| Sensitivity: add abscess or deep fluid | 3136/8533 | 36.8 (35.7-37.8) |
| Sensitivity: primary-diagnosis episodes only | 1413/3170 | 44.6 (42.8-46.3) |
| Sensitivity: excluding 97 dual-coded (PJI + osteomyelitis) episodes | 2617/7447 | 35.1 (34.1-36.2) |
| Prosthetic joint infection only | 1337/2749 | 48.6 (46.8-50.5) |
| Native osteomyelitis only | 1020/3829 | 26.6 (25.2-28.1) |

The no-growth fraction was stable across specimen-definition and case-definition sensitivity

analyses. The patient-clustered bootstrap 95% CI for the primary specimen-level fraction (34.0%-37.3%) was slightly wider than the naive interval (34.6%-36.7%), as expected given within-patient correlation.

#### Table S 7. Full antimicrobial susceptibility

| Antimicrobial agent | Susceptible, n | Intermediate, n | Resistant, n | Isolates tested, n | Resistant, % |
| --- | --- | --- | --- | --- | --- |
| Gentamicin | 5,085 | 167 | 430 | 5,682 | 7.6 |
| Trimethoprim-sulfamethoxazole | 3,798 | 1 | 548 | 4,347 | 12.6 |
| Erythromycin | 1,418 | 15 | 2,296 | 3,729 | 61.6 |
| Levofloxacin | 2,164 | 22 | 1,513 | 3,699 | 40.9 |
| Oxacillin | 1,709 | 0 | 1,767 | 3,476 | 50.8 |
| Clindamycin | 2,044 | 22 | 1,403 | 3,469 | 40.4 |
| Tetracycline | 2,717 | 13 | 423 | 3,153 | 13.4 |
| Vancomycin | 2,769 | 4 | 321 | 3,094 | 10.4 |
| Ciprofloxacin | 1,393 | 80 | 703 | 2,176 | 32.3 |
| Ceftazidime | 1,636 | 135 | 401 | 2,172 | 18.5 |
| Tobramycin | 1,801 | 144 | 188 | 2,133 | 8.8 |
| Cefepime | 1,679 | 128 | 323 | 2,130 | 15.2 |
| Meropenem | 1,850 | 68 | 187 | 2,105 | 8.9 |
| Rifampin | 1,853 | 5 | 68 | 1,926 | 3.5 |
| Piperacillin-tazobactam | 1,568 | 90 | 206 | 1,864 | 11.1 |
| Ceftriaxone | 1,150 | 33 | 331 | 1,514 | 21.9 |
| Ampicillin | 730 | 10 | 566 | 1,306 | 43.3 |
| Penicillin G | 621 | 29 | 455 | 1,105 | 41.2 |
| Ampicillin-sulbactam | 518 | 113 | 272 | 903 | 30.1 |
| Cefazolin | 384 | 8 | 317 | 709 | 44.7 |
| Linezolid | 540 | 1 | 0 | 541 | 0.0 |
| Daptomycin | 528 | 1 | 4 | 533 | 0.8 |
| Nitrofurantoin | 190 | 80 | 84 | 354 | 23.7 |
| Amikacin | 215 | 10 | 40 | 265 | 15.1 |
| Piperacillin | 150 | 7 | 65 | 222 | 29.3 |
| Cefuroxime | 78 | 9 | 15 | 102 | 14.7 |
| Imipenem | 68 | 16 | 15 | 99 | 15.2 |
| Minocycline | 11 | 0 | 0 | 11 | 0.0 |
| Ertapenem | 10 | 0 | 0 | 10 | 0.0 |
| Doxycycline | 2 | 0 | 5 | 7 | 71.4 |
| Fluconazole | 4 | 0 | 0 | 4 | 0.0 |
| Imipenem-relebactam | 3 | 0 | 0 | 3 | 0.0 |
| Ceftazidime-avibactam | 2 | 0 | 0 | 2 | 0.0 |
| Caspofungin | 1 | 0 | 0 | 1 | 0.0 |
| Aztreonam | 0 | 0 | 1 | 1 | 100.0 |
| Ceftolozane-tazobactam | 1 | 0 | 0 | 1 | 0.0 |
| Omadacycline | 1 | 0 | 0 | 1 | 0.0 |

*Susceptible / intermediate / resistant counts by antimicrobial agent among deep musculoskeletal isolates with an interpretable result, pooled across all organisms. Resistant % is R / (S + I + R). Agents are ordered by number of isolates tested. Because intrinsic resistance varies by organism, these pooled per-agent percentages are not organism-specific and differ from the organism-stratified rates in Table 3 — for example, pooled oxacillin resistance exceeds the S. aureus-specific value because coagulase-negative staphylococci are included. Table 3 should be used for organism-specific interpretation.*

#### Table S 8. Analytic flow

| Step | Episodes (patients) | Deep MSK culture specimens |
| --- | --- | --- |
| Code-defined orthopedic-infection episodes | 7697 (4358) | — |
| ... prosthetic joint infection (ICD-9 996.66 / ICD-10 T84.5x) | 1089 | — |
| ... native osteomyelitis | 5715 | — |
| ... other internal-device infection (ICD-9 996.67 / ICD-10 T84.6-T84.7) | 893 | — |
| Episodes with >=1 deep MSK culture (primary analytic cohort) | 3560 (2603) | 7700 |
| Culture-positive episodes (>=1 deep isolate) | 2797 | — |

Episodes without a deep musculoskeletal culture were excluded from the organism-spectrum and no-growth analyses; this excluded group may include medically managed infections and miscoded episodes, a limitation noted in the main text.

#### Fig. S 1

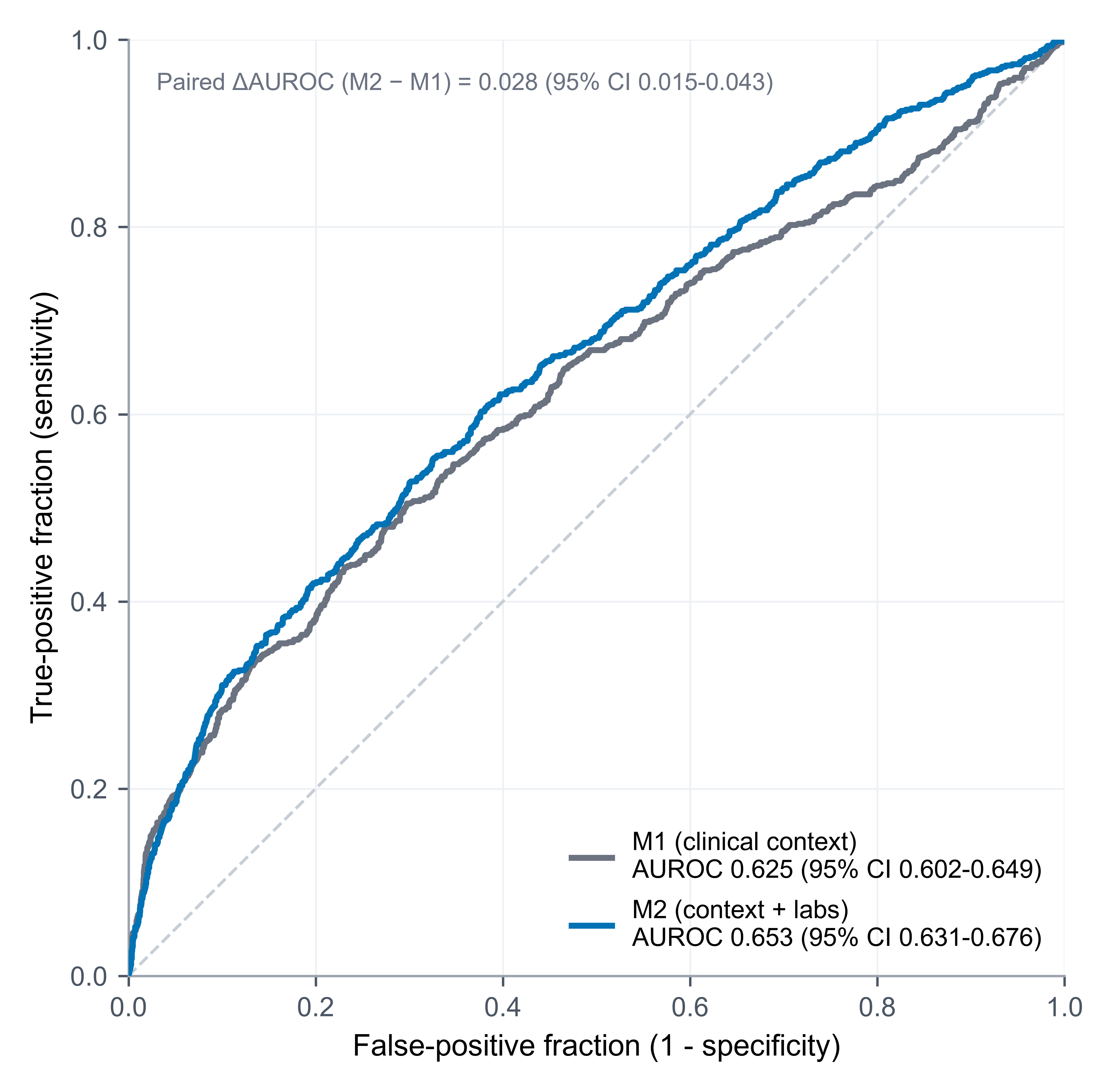

*Fig. S 1. Out-of-fold receiver operating characteristic curves for the anticipatability probe. Model M1 uses clinical-context features only (infection type, deep source-category flags, age, sex, race group, insurance); model M2 adds inflammatory and nutritional laboratory markers. Curves are pooled out-of-fold predictions from 5-fold stratified, patient-grouped cross-validation (no patient split across folds); 95% CIs are from 2000-sample bootstrapping. The modest discrimination, barely improved by laboratory markers, is consistent with deep-specimen no-growth not being an artifact readily explained by routine structured data.*
