## Supplementary material for "Organism spectrum and no-growth fraction of deep specimens in code-defined orthopedic infection: a reproducible, cross-sectional MIMIC-IV benchmark": STROBE-RECORD reporting checklist

**STROBE and RECORD reporting checklist**

This checklist combines the STROBE statement for cross-sectional studies with the RECORD extension for studies conducted using routinely collected health data. The Location column references the relevant section of the manuscript and the Supplement (eMethods, eTables, eFigure); page numbers are assigned at typesetting.

| **Item** | **Recommendation** | **Location in manuscript / Supplement** |
| --- | --- | --- |
| **Title and abstract** | | |
| **1** | (a) Study design in title or abstract. (b) Informative and balanced abstract summary. | Title; Abstract (Design; Structured summary) |
| **Introduction** | | |
| **2** | Background/rationale for the investigation. | Introduction |
| **3** | State specific objectives, including any prespecified hypotheses. | Introduction (final paragraph; Aims 1-5) |
| **Methods** | | |
| **4** | Present key elements of study design early in the paper. | Methods, Data source and reporting; eMethods 1-2 |
| **5** | Setting, locations, and relevant dates (recruitment, exposure, follow-up, data collection). | Methods, Data source and reporting; eMethods 1 (MIMIC-IV v3.1, 2008-2022 admissions) |
| **6** | (a) Eligibility criteria, sources, and methods of selection of participants. | Methods, Cohort; eMethods 2; eTable 8 (analytic flow) |
| **RECORD 6.1** | Detail methods of study population selection (codes/algorithms used to identify the population). | eMethods 2 (ICD-9/ICD-10 code prefixes); eTable 8 |
| **RECORD 6.2** | Validation of the codes/algorithms used to select the population. | eMethods 2 and Discussion, Limitations (codes not clinically validated; cohort framed as code-defined) |
| **RECORD 6.3** | If validation was conducted, describe population used and validation results. | Not applicable (no code validation performed; stated as a limitation) |
| **7** | Clearly define all outcomes, exposures, predictors, potential confounders, effect modifiers; give diagnostic criteria. | Methods, Measures; eMethods 3-4 (specimen taxonomy, organism dictionary, measure definitions) |
| **RECORD 7.1** | Complete list of codes and algorithms used to classify exposures, outcomes, confounders, and effect modifiers. | eMethods 3; eTable 2 (specimen taxonomy); eTable 3 (organism dictionary); code/spec_map.py, code/org_map.py |
| **8** | For each variable, give sources of data and details of methods of assessment. | Methods, Measures; eMethods 3-4 |
| **9** | Describe any efforts to address potential sources of bias. | Methods, Statistical analysis; Discussion, Limitations (sampling, coding, informative missingness) |
| **10** | Explain how the study size was arrived at. | Methods, Cohort; eTable 8 (all code-defined episodes with >=1 deep culture) |
| **11** | Explain how quantitative variables were handled in the analyses. | Methods, Statistical analysis; eMethods 4-5; Model card (eMethods 6) |
| **12** | (a) Statistical methods including confounding control. (b) Subgroups/interactions. (c) Missing data. (d) Sampling strategy. (e) Sensitivity analyses. | Methods, Statistical analysis; eMethods 5; eTable 6 (sensitivity analyses) |
| **RECORD 12.1** | Detail methods of data cleaning and linkage of databases. | eMethods 1-3 (hosp and icu modules linked on hadm_id/subject_id; normalization dictionaries) |
| **RECORD 12.2** | State whether the study included person-level, institutional-level, or other data linkage. | eMethods 1 (single-center person-level EHR data; internal module linkage only) |
| **RECORD 12.3** | Access to and cleaning of the population/data source available to investigators. | eMethods 1; Data availability (credentialed PhysioNet access; DATA_ACCESS.md in repository) |
| **Results** | | |
| **13** | (a) Numbers of individuals at each stage. (b) Reasons for non-participation. (c) Flow diagram. | Results, Cohort; eTable 8 (analytic flow) |
| **RECORD 13.1** | Detail selection of persons included/excluded; justify inclusion/exclusion when persons excluded. | eTable 8; Discussion, Limitations (episodes without a deep culture excluded from spectrum/no-growth aims) |
| **14** | (a) Characteristics of study participants and information on exposures/confounders. (b) Missing data per variable. | Results, Cohort; Table 1; Model card (eMethods 6) marker missingness |
| **15** | Report numbers of outcome events or summary measures. | Results, Aims 1-5; Tables 2-3; eTables 4-7 |
| **16** | (a) Unadjusted and confounder-adjusted estimates with 95% CIs. (b) Category boundaries. (c) Absolute risk if relevant. | Results, Aims 2 and 5; Table 3; eMethods 5 |
| **17** | Report other analyses done (subgroups, interactions, sensitivity analyses). | Results, Aims 2-5; eTables 5-6; eFigure 1 |
| **Discussion** | | |
| **18** | Summarize key results with reference to study objectives. | Discussion (opening) |
| **19** | Discuss limitations, sources of potential bias or imprecision, direction and magnitude of bias. | Discussion, Limitations |
| **RECORD 19.1** | Discuss the implications of using data not created or collected to answer the research question(s); misclassification, unmeasured confounding, missing data, changing eligibility over time. | Discussion, Limitations (administrative coding, unmeasured severity and outpatient antibiotics, informative missingness) |
| **20** | Give a cautious overall interpretation considering objectives, limitations, and other evidence. | Discussion; Conclusions |
| **21** | Discuss the generalizability (external validity) of the study results. | Discussion, Limitations (single US academic center; no external validation) |
| **Other information** | | |
| **22** | Give the source of funding and the role of funders. | Funding statement |
| **RECORD 22.1** | State whether investigators had access to the database population used to create the study population, and provide information on how the database can be accessed. | Data availability; Code availability (credentialed PhysioNet access; DATA_ACCESS.md) |

*STROBE: von Elm E, et al. Ann Intern Med. 2007;147(8):573-577. RECORD: Benchimol EI, et al. PLoS Med. 2015;12(10):e1001885.*
